# Circulating blood circular RNA in Parkinson’s Disease; a systematic study

**DOI:** 10.1101/2024.01.22.24301623

**Authors:** Aleksandra Beric, Yichen Sun, Santiago Sanchez, Charissa Martin, Tyler Powell, Jose Adrian Pardo, Jessie Sanford, Juan A. Botia, Carlos Cruchaga, Laura Ibanez

**Affiliations:** Department of Psychiatry, Washington University in Saint Louis School of Medicine; NeuroGenomics and Informatics Center, Washington University in Saint Louis School of Medicine; Departamento de Ingeniería de la Información y las Comunicaciones; Universidad de Murcia, Murcia, Spain; Department of Neurodegenerative Diseases, Institute of Neurology, University College London, London UK; Department of Neurology, Washington University in Saint Louis School of Medicine; Hope Center for Neurological Disorders, Washington University in Saint Louis School of Medicine; The Charles F. and Joanne Knight Alzheimer Disease Research Center, Washington University in Saint Louis; Department of Genetics, Washington University in Saint Louis School of Medicine

**Author notes:** Correspondence: Laura Ibanez, PhD Washington University in Saint Louis School of Medicine 4444 Forest Park Ave Campus Box 8134 Saint Louis, MO 63110.

## Abstract

We aimed to identify circRNAs associated with Parkinson’s disease (PD) by leveraging 1,848 participants and 1,789 circRNA from two of the largest publicly available studies with longitudinal clinical and blood transcriptomic data. To comprehensively understand changes in circRNAs we performed a cross-sectional study utilizing the last visit of each participant, and a longitudinal (mix model) analysis that included 1,166 participants with at least two time points. We identified 192 circRNAs differentially expressed in PD participants compared to healthy controls, with effects that were consistent in the mixed models, mutation carriers, and diverse ancestry. Finally, we included the 149 circRNA in a model with a ROC AUC of 0.825, showing that have the potential to aid the diagnosis of PD. Overall, we demonstrated that circRNAs play an important role in PD and can be leveraged as biomarkers.

## BACKGROUND

Parkinson’s disease (PD) affects more than six million people worldwide, with a prevalence projected to double in the next decades.^1^ It is a neurodegenerative disease clinically defined by resting tremor, rigidity, bradykinesia, and postural instability.^2^ Pathologically, it is characterized by Lewy bodies (LB) and neurites that are composed by aggregated and phosphorylated alpha-synuclein (α-Syn) and the degeneration of substantia nigra. Its cause is not fully understood yet. It is known that there are both genetic and environmental risk factors, but the complete causal pathway, or pathways, still remain elusive.

Circular RNAs (circRNAs) are non-coding RNAs that are the result of backsplicing events that take place during the maturation of linear RNA and lead to the creation of a covalently closed loop and an increase in their stability.^3–6^ They are highly expressed in the nervous system, especially in synapses^7^ and have a role in neuronal development and aging.^5^ Even though their exact function is still to be deciphered, many functions have been already attributed to circRNAs. They have been found to act as miRNA sponges to regulate gene expression, interact with proteins, and to generate protein products among many others.^3,4,7–12^ Additionally, they are differentially accumulated in several pathological states, including that of the central nervous system diseases^3,4,7–12^, such as PD. In healthy conditions, the substantia nigra accumulates circRNA in an age-related manner. That accumulation is lost in PD, with a reduction on the overall number of circRNAs.^11^ The same study by Hanan *et al.* identified 23 circRNA differentially expressed in not only substantia nigra, but also medial temporal gyrus and amygdala from PD affected brains compared to controls.^11^ In blood, a study comparing four PD cases and four controls described 129 circRNAs up-regulated, and 282 down-regulated^13^ in PD participants compared to controls. The circRNA host genes of the identified circRNAs were enriched in PD terms according to the Kyoto Encyclopedia of Genes and Genomes (KEGG). More recently, a high throughput study found three circRNAs downregulated in PD blood. Unfortunately, they were unable to replicate the results in an independent cohort.^14^ A study analyzing 87 circRNA from Peripheral Blood Mononuclear Cells (PBMCs) in 60 PD participants and 60 controls identified six circRNA downregulated in PD.^10^ Finally, they used four of them to build a classifier that showed an Area Under the ROC curve (AUC) of 0.86, which adds evidence to the potential leverage of circular RNAs as biomarkers for neurodegenerative diseases.

Currently, PD is diagnosed based on clinical and neuroimaging criteria, and then monitored using clinical tests that assess the motor and non-motor symptoms of the disease. However, there are no molecular diagnostic or prognostic biomarkers available for PD, or in general, for most of the neurodegenerative diseases.^15^ Circular RNA are highly stable and very abundant in the brain, which means they can potentially leak to the CSF or blood via blood brain barrier breakdown.^16^ If informative, circRNA can be measured by real-time PCR, are stable due to being circular, and can be measured in blood, qualities highly desired in biomarker development.^6,15^

To date, there is one high-throughput screening of circRNAs in blood of PD individuals, which focused on early-stage PD.^14^ Outside of the high-throughput realm, there are targeted analyses using PBMC with promising results.^10^0 In here, we have quantified circRNAs in blood from the two largest longitudinal studies, the national Institute of Neurological Disorders and Parkinson’s Disease Biomarkers Program (PDBP)^17^, and the Parkinson’s Progression Markers Initiative (PPMI)^18^, to describe the landscape of circRNAs in blood of PD participants compared to controls and evaluate their value as diagnostic and prognostic biomarkers.

## MATERIAL & METHODS

### Study Design

We accessed the largest PD blood RNAseq datasets publicly available to date from the PDBP and PPMI studies. After raw data processing and stringent quality control, we compared the circular transcriptome between European Ancestry PD participants and controls to identify differentially abundant circRNAs. We leveraged the PDBP dataset (N=1,177) for discovery and PPMI (N=671) for replication, followed by meta-analyses. We used gene-collapsed circRNA counts in the analyses and corrected all p-values using Benjamini-Hochberg (FDR) correction. Then, we investigated if the identified circRNAs were also associated with disease progression by performing longitudinal analyses of the RNAseq data collected across five visits. To confirm that the findings were not due to differences in cell proportions, or originated from the linear transcriptome, we performed the same analyses including cell proportions calculated using digital deconvolution, and on the linear transcriptome. Then we investigated if the same circRNAs were differentially abundant in the African Ancestry individuals available in PDBP and PPMI. To assess if changes in circRNA accumulation can be observed prior to symptom onset, and thus if circRNA had a potential to be used as early stage biomarkers, we evaluated the trajectory of the identified circRNAs in individuals at high risk of PD (known PD-related mutation carriers and participants with REM sleep Behavior Disorder (RBD) or hyposmia). We evaluated if the most promising circRNAs, whose accumulation wasn’t affected by cellular composition or medication, associated with disease severity. To do so, we tested their association with the Unified Parkinson’s Disease Rating Scale (UPDRS) Part III or motor examination (UPDRS-III), and cognitive status measured by The Montreal Cognitive Assessment (MoCA). Additionally, and to understand the biology of the identified circRNAs, we performed data integration with microRNA quantification (miRNA) and *in-silico* functional analyses. And finally, to assess their diagnostic capacity in early stages of the disease, we developed machine learning models to identify those circRNAs more relevant in the prediction task. A summary of the study design can be found in Figure 1.

**Figure 1.**
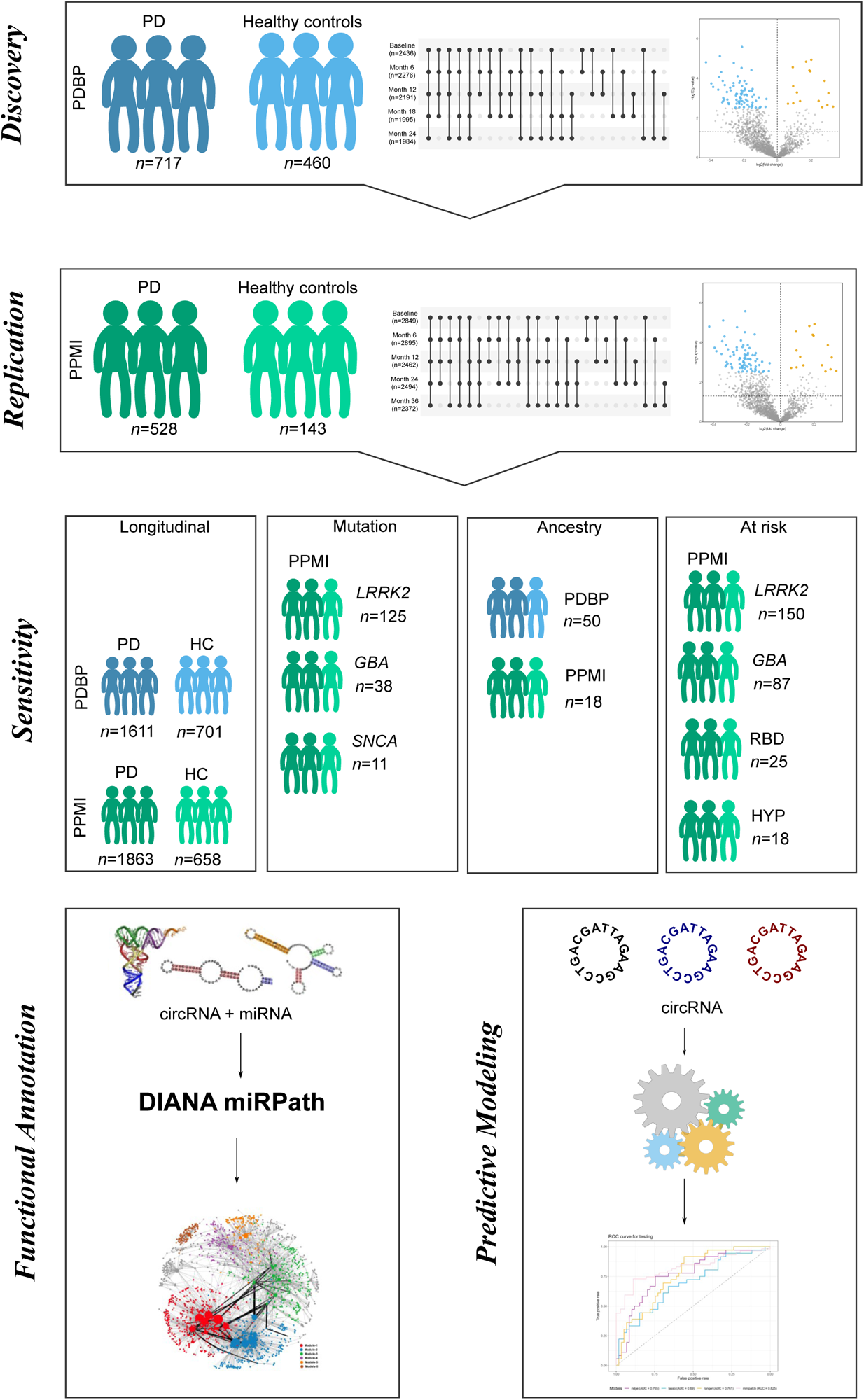
Study design summary. We have followed a two-stage cross-sectional analysis with discovery (PDBP) and replication (PPMI) phases. Then we have performed sensitivity analysis for the top hits by approaching the analysis in a longitudinal manner (Mixed model), stratifying by mutation, exploring individuals from African Ancestry, and those at risk due to being a carrier of a PD-causing mutation, or suffering from RBD or hyposmia. Finally, we have performed functional annotation via circRNA-miRNA integration and leveraged th e circRNA to build predictive models.

### Dataset Description

The present study includes two independent and publicly available datasets with longitudinal clinical and transcriptomic data available: PDBP^17^ and PPMI^18^. Both are observational and multi-center studies aimed at identifying biomarkers of PD progression and improving the understanding of PD pathobiology. Participants (N=1,848; Table 1) are followed longitudinally with clinical assessments, imaging, and biospecimen collection every six months. To assess the accumulation of circRNA, we maximized our dataset and clinical differences by selecting the last assessment of each participant. We have included a total of 717 cases and 460 control participants of European descent with RNAseq data available from the PDBP study (Table 1) as discovery, and 528 cases and 143 control participants of European descent form the PPMI study (Table 1) to identify circRNA differentially accumulated in the blood of cases compared to controls. Unsurprisingly, both datasets had slightly higher proportion of male participants (>60%), with the exception of healthy control participants in PDBP, with more than 50% of females. Further, PDBP and PPMI participants are similar in terms of symptom severity as measured by UPDRS-III and MoCA. Regarding UPDRS-III scale, 75% of healthy controls present scores between 0-2, whereas 75% of symptomatic participants score between 16-33 as expected. MoCA scores vary less between healthy controls and symptomatic participants, with a narrow range of means between 25-27 (Table1). This is probably due to the enrolment of recently diagnosed individuals in both studies, in other words, individuals at the beginning of the disease which may or may not develop dementia in the future. We have leveraged the participants with African descent (N=68) from the PDBP (N=50) and PPMI (N=18) studies to investigate if the findings are ancestry independent (Supplemental Table 1).

**Table 1.** Demographic characteristics corresponding to the European descent individuals included in the cross-sectional analyses.

|  | Parkinson's Disease Biomarker Program (PDBP) |  | Parkinson's Progression Markers Initiative (PPMI) |  |
| --- | --- | --- | --- | --- |
|  | Healthy Controls | Parkinson's Disease | Healthy Controls | Parkinson's Disease |
| **Participants (N)** | 460 | 717 | 143 | 528 |
| **Mean age (IQR)** | 62.83<br>(56.00-71.00) | 65.85<br>(60.00-72.00) | 63.93<br>(59.00-71.00) | 64.33<br>(57.75-71.00) |
| **Female (N, %)** | 251<br>(54.56%) | 269<br>(37.52%) | 52<br>(36.36%) | 203<br>(38.45%) |
| **Mean UPDRS-III (IQR)** | 2.13<br>(0.00-2.00) | 25.88<br>(16.00-33.00) | 1.46<br>(0.00-2.00) | 24.95<br>(15.75-33.25) |
| **Mean MOCA (IQR)** | 26.61<br>(25.00-28.50) | 25.15<br>(23.00-28.00) | 27.34<br>(26.00-29.00) | 26.32<br>(25.00-29.00) |

N=Sample Size; IQR=Interquartile Range; UPDRS-III=Unified Parkinson's Disease Rating Scale Part III; MoCA=Montreal Cognitive Assessment

Longitudinal RNAseq data was available for 1,846 unique participants across five visits in both PDBP and PPMI studies (Table 2). Visits in PDBP were six months apart, while PPMI participants were followed every 6 months during the first year, and every twelve months thereafter. Mean age of PDBP participants with more than one visit is greater than that of PPMI participants. PDBP cohort is more evenly distributed between sexes, with 40-50% of female participant across visits, while PPMI consists of 30-40% female participants. PPMI dataset also consisted of participants with PD-associated mutations and participants with risk-related PD syndromes, RBD (N= 25) or hyposmia (N=18) (Supplemental Table 2). *LRRK2* mutations were the most prevalent in both symptomatic (N=125) and at risk (N=150) participants; more than 50% were female, with mean age >60, similar to sporadic PD participants (Supplemental Table 2). In contrast, *SNCA* mutation carriers were the least prevalent (N=11 symptomatic and N=3 at risk participants) with mean age <55, notably lower than other participants (Supplemental Table 2).

**Table 2.** Demographic characterizing of the European descent participants included in the longitudinal analysis by time of sample collection. Each participant must have available data in at least two time-points.

| Sampling time | Group | Parkinson's Disease Biomarker Program (PDBP) |  |  | Parkinson's Progression Markers Initiative (PPMI) |  |  |
| --- | --- | --- | --- | --- | --- | --- | --- |
|  |  | Samples (N) | Mean age (IQR) | Female (N, %) | Samples (N) | Mean age (IQR) | Female (N, %) |
| Baseline | HC | 166 | 63.95<br>(56.00-71.00) | 80<br>(48.19) | 143 | 61.12<br>(56.00-68.00) | 52<br>(36.36) |
|  | PD | 362 | 64.77<br>(59.00-71.00) | 147<br>(40.60) | 474 | 61.54<br>(55.00-69.00) | 183<br>(38.60) |
| Month 6 | HC | 139 | 64.59<br>(56.50-71.00) | 66<br>(47.48) | 129 | 61.64<br>(57.00-69.00) | 48<br>(37.21) |
|  | PD | 337 | 65.45<br>(59.50-71.50) | 140<br>(41.54) | 341 | 62.41<br>(55.00-69.00) | 139<br>(40.76) |
| Month 12 | HC | 146 | 64.89<br>(57.00-71.75) | 68<br>(46.57) | 130 | 62.19<br>(57.00-69.00) | 45<br>(34.61) |
|  | PD | 333 | 65.94 (60.00-72.00) | 135<br>(40.54) | 388 | 62.34<br>(56.00-69.00) | 150<br>(38.66) |
| Month 18 | HC | 128 | 64.48<br>(57.50-70.50) | 64<br>(50.00) | - | - | - |
|  | PD | 299 | 66.43<br>(60.50-72.50) | 124<br>(41.47) | - | - | - |
| Month 24 | HC | 122 | 66.01<br>(59.00-72.00) | 60<br>(49.18) | 129 | 62.88<br>(58.00-70.00) | 49<br>(37.98) |
|  | PD | 280 | 67.17<br>(61.00-73.00) | 124<br>(44.28) | 381 | 63.74<br>(57.00-71.00) | 142<br>(37.27) |
| Month 36 | PD | - | - | - | 127 | 63.88<br>(59.00-71.50) | 48<br>(37.79) |
|  | HC | - | - | - | 279 | 64.54<br>(58.00-72.00) | 94<br>(33.69) |

N=Sample Size; IQR=Interquartile Range; HC= Healthy Control; PD=Parkinson's Disease

### Data Processing and Quality Control

We accessed raw transcriptomic data from a total of 6,362 ribodepleted blood RNA samples from 1,848 unique participants from the PDBP and PPMI studies. Data generation, processing, and quality control for these datasets have been described elsewhere (REF). Due to the lack of availability of circular counts, we re-processed the raw files using our inhouse pipelines to obtain linear and circular counts (REF). For linear counts, we followed the TOPMed pipline using the GRCh38 genome reference and the GENCODE 33 annotation^7,19^ (https://github.com/broadinstitute/gtex-pipeline/blob/master/TOPMed_RNAseq_pipeline.md). Briefly, the raw reads were aligned to the human reference genome using STAR (v.2.7.1a)^20^ and alignment quality was evaluated using sequencing metrics calculated using Picard tools (v.2.8.2).^21^ Gene expression was quantified using Salmon (v.1.2.0).^22^ All transcripts or genes with less than ten reads in more than 90% of the individuals were removed from the analyses. All transcripts were collapsed to gene level for analysis.

Circular RNA detection, annotation and quantification was performed using Detect CircRNA from Chimeric Reads (DCC v.0.4.8)^23^ using the software developer guidelines and as described previously.^24^ Shortly, the raw reads were aligned a second time to the human reference genome (GRCh38) using STAR^20^ in chimeric alignment mode. Similar to linear RNA, circRNAs were collapsed by host gene prior to analyses. CircRNAs with missing counts in more than 75% of the samples were removed from the analyses. We processed the samples from each visit and study separately. Finally, we integrated the circRNA counts for each study (Supplementary Figure 1A and 1B). For each study separately, we calculated Principal Component Analysis (PCA) using the 500 most variable circular transcripts and removed samples that were more than three-standard deviations from the mean of PC1 or PC2 (Supplemental Figure1C and 1D). To normalize the counts, we used DESeq2^25^ to adjust for library complexity and variance stabilizing transformation (vst) to obtain the final count matrix. As an additional QC step, we calculated count ratios between circRNAs and linear RNAs and kept only those circRNAs that had circRNA:linear RNA ratio of at least 0.1 in three or more samples.^7^ Only circular transcripts that passed QC in both PDBP and PPMI datasets were included in the analyses (N=1,789).

### Cross-Sectional Differential Abundance Analysis

We performed cross-sectional differential abundance analyses using DESeq2.^25^ To maximize the detection of differentially abundant circular transcripts due to PD and taking into consideration the longitudinal nature of PDBP and PPMI, we included one sample per individual, corresponding to the most recent visit, regardless of mutation status. No at-risk individuals were included in this analysis. All analyses were adjusted by sex and age at draw. Following discovery in the PDBP and replication in the PPMI dataset, we meta-analyzed the results using the R package metaRNAseq.^26^ Only circRNAs with same direction of effect in both PDBP and PPMI datasets were considered for meta-analyses. All p-values were multiple test corrected using the FDR correction. FDR p-values below 0.05 were considered significant.

To ensure the robustness of the results, and that they were not driven by differences in cellular composition, EPIC^27^ was used to obtain the cell proportions, and those included in the model. Similarly, to evaluate if the findings could be driven by changes on the linear forms of the host genes, we performed the same analyses described above including the linear counts. Medication can be another confounding factor, thus we wanted to understand if PD medication (L-Dopa or Dopamine Agonist) had any impact on the differential accumulation. In consequence, and similar to what we did for cellular composition and linear counts, we adjusted the analyses by the participants medication status (yes or no) and evaluated if the association was still significant. Medication information is difficult to obtain and harmonize. We had access to individual data for the PPMI study only, that we curated manually.

### Cross-Sectional Sensitivity Analyses

To better understand the role of the identified circRNA in the disease, we assessed whether the counts of the circular transcripts identified in the cross-sectional analyses were correlated with UPDRS III or MoCA for those participants with the data available. Finally, to investigate if there was any difference between sporadic PD and mutation carriers in regard to circRNA abundance, we performed sensitivity analyses by dividing the population is smaller groups (*LRRK2*, *GBA*, or *SNCA* mutation carriers) and compared the circRNA counts to those of healthy controls or non-mutation carriers. Additionally, we leveraged data from at risk participants (PPMI participants that carry a known PD mutation but have not been diagnosed with PD or that suffer from RBD or hyposmia) to test if the significant circRNAs were differentially accumulated in early stages of the disease compared to healthy controls.

To leverage the diversity that these datasets include, we performed circRNA differential expression analyses, focusing on the findings from the cross-sectional differential abundance analysis, in participants of African ancestry following the methodology described above. Given the limited participants available, and to maximize our statistical power, we combined participant data that passed QC from both PDBP and PPMI datasets (N=14 PD participants and N=17 healthy controls).

### Longitudinal Differential Abundance Analysis

To harness the longitudinal characteristics of the two studies included in this manuscript we use mixed models to perform differential abundance analyses. We included participants from PDBP (N=547) and PPMI (N=619) datasets with at least two clinical visits and RNAseq available after QC regardless of mutation status. We modeled the trajectories using linear mixed model with *counts×time* as the interaction term. All analyses were adjusted by circular transcript counts at first visit, sex, and age at draw. Participant ID was used as random effect. The overall approach was similar to the one described above but focusing on the circular transcripts that were found significant in the cross-sectional analyses. Briefly, PDBP and PPMI were tested separately to perform a subsequent meta-analysis using only European ancestry cases and controls. Then, we explored if the linear transcripts, the cell proportion, or medication had an influence on the results. We investigated if the longitudinal circRNA counts for each of the transcripts was correlated with disease severity progression measured by UPDRS III and MoCA. Unfortunately, the number of African ancestry individuals with at least two visits was very limited (N=16), thus no testing on diverse ancestry was performed. All p-values were FDR adjusted based on the number of circular transcripts identified in the cross-sectional.

### *In-Silico* Functional Study

We explored the biological implications of the identified circRNA accessing the Circular RNA Interactome website^28^ (accessed July 2023, last database update January 30^th^ 2020) to predict which miRNAs have the potential to be targeted by each of the circRNA species identified in the present study. To reduce the number of miRNA, and since miRNA sequence data is available for the PPMI dataset, we explored if the counts of any of the predicted miRNA were correlated with the circRNA counts to identify the ones that are more likely to have biological consequences. After accessing the available miRNA count data from the PPMI study and filtering the low count miRNA data (standard quality control parameter of at least five counts in 90% of the sample). The miRNA counts were normalized using DESeq2^25^ and the vst function (similar to what was described for the circular and the linear RNAs). We used Pearson correlation to assess which of the predictions hold true at biological level. With the list of most significant miRNAs correlated with each circRNA, we performed pathway analyses, grouped by circRNA, using the microT-CDS algorithm and the Kyoto Encyclopedia of Genes and Genomes (KEGG) form the DIANA mirPath software version 3^29^, to identify pathways regulated by the miRNAs and, consequently, the circRNAs targeting them.

### Predictive model development and evaluation

We tested whether circRNA levels can be used to differentiate between early-stage PD and healthy controls. To do so, we focused on samples from the first visit in PDBP (N= 528) and PPMI (N=617) datasets, and 2,849 circRNAs shared between the two datasets in the first visit. We employed several linear and non-linear machine learning approaches: L1 and L2 regularization as implemented in the glmnet R package^30,31^, random forests from the Ranger R package^32^, and the MiniPatch learning algorithm.^33^ Since L1 regularization, Ranger and MiniPatch algorithms have a built-in feature selection, the whole set of 2,849 circRNAs was used as input for all these methods. L2 regularization does not perform any internal feature selection, so we supplied it with counts for the best performing L1 regularization, Ranger and MiniPatch models. Hyperparameters of Ranger and MiniPatch were optimized by using a grid search over a subset of possible hyperparameter values. The best model was selected based on the best ROC-AUC and testing balanced accuracy out of 10 repeated 10-fold cross-validation experiments.

## RESULTS

### CircRNA are differentially accumulated in blood of PD cases compared to controls

To identify circRNA differentially accumulated in the blood of PD cases compared to controls, we used a cross-sectional approach, selecting the most recent blood sample available per individual. We included 1,177 PDBP participants as discovery (N_cases_=717; N_controls_=460) and 671 from PPMI as replication (N_cases_=528; N_controls_=143; Table1, Figure1). Meta-analyses revealed 192 circRNAs to be differentially accumulated when comparing cases and controls after multiple test correction (Figure 2A, Supplemental Table 3). Of those, 71 were nominally significant in both the discovery and the replication dataset. Moreover, nine were significant after multiple test correction in both datasets (Figure 2B), on which we focused for downstream analyses.

**Figure 2.**
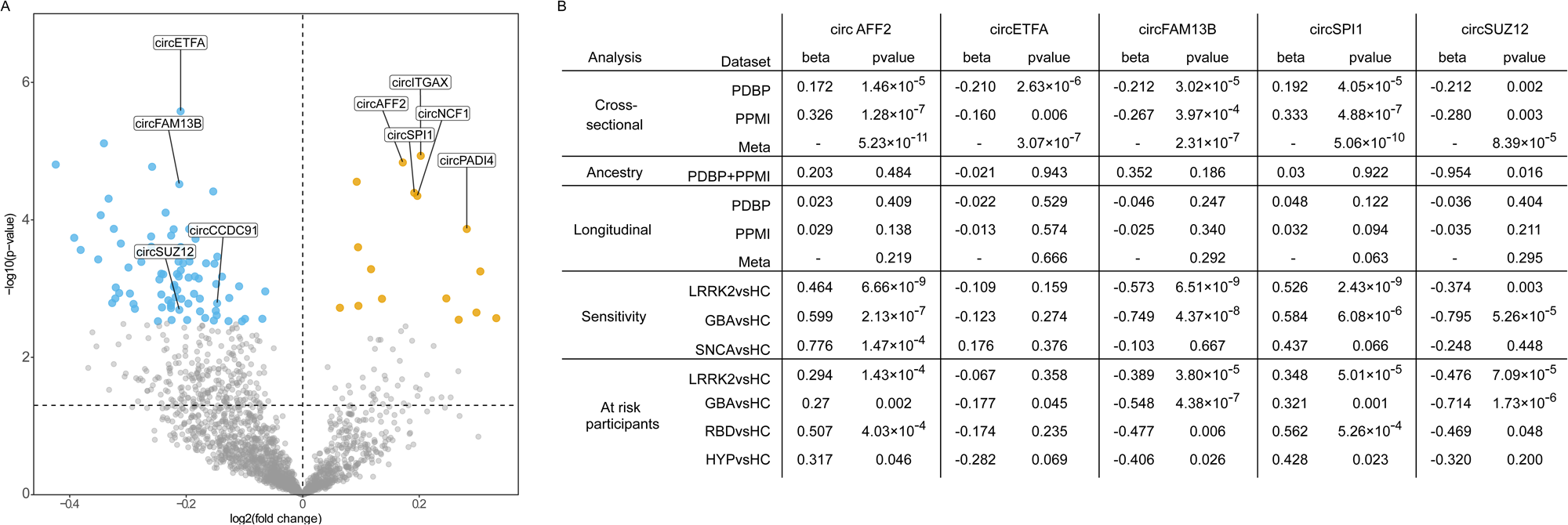
Results of the cross sectional analysis; A. Volcano plot showing circRNAs that are differentially expressed in the PDBP dataset, with replicated circRNAs in PPMI labeled; B. Summary results of the main findings including sensitivity analyses

Since we are analyzing whole blood trasncriptome, we repeated the analysis including cell proportions. Only circ*CCDC91* (log_2_FC=0.263, p=0.253; Supplemental Table 4), was associated with differences in cellular composition. This association was driven by neutrophils (p<2×10^-16^), CD4+ T-cells (1.76×10^-10^) and monocytes (p=9.54×10^-7^). Medication is another key component that might be affecting the transcriptomic landscape. Even though medication is collected in both datasets, detailed and comprehensive medication information was only available for PPMI. The association of three circRNAs, circ*ITGAX* (log_2_FC=0.062, p=0.515), circ*PADI4* (log_2_FC=0.177, p=0.231), and circ*NCF1* (log_2_FC=0.183, p=0.095) (Supplemental Figure 2; Supplemental Table 4) seem to be driven by the presence of medication and not the disease *per se*. Finally, this resulted in a total of five high-confidence circRNAs, circ*AFF2*, circ*ETFA*, circ*FAM13B*, circ*SPI1* and circ*SUZ12* (Figure 2, Supplemental Table 4), that were followed up in further analyses.

Next, we verified that linear RNA counts might be driving our results by evaluating their differential accumulation using the same model. None of them was significant, except for *FAM13B*. We observed decreased levels of circ*FAM13B* in PD (log_2_FC=-0.212, p=3.97×10^-4^), but as previously described^13^, we found that the linear form of *FAM13B* (log_2_FC=0.036, p=3.99×10^-5^) was also differentially accumulated in PD cases compared to controls. To understand if the signal was driven by the linear form of *FAM13B*, we included in the analyses both the linear and the circular RNA counts in addition to other covariates. Both forms of *FAM13B*, linear (log_2_FC=0.043, p=5.73×10^-6^) and circular (log_2_FC=-0.201, p=4.35×10^-7^), remained significant, suggesting that both participate in the association. The remaining linear RNA were not significantly associated with PD in their linear forms (Supplemental Table 5).

Finally, we explored the effect that diverse ancestry has on circRNA accumulation and found that none of the five circRNAs were statistically significant in the comparison. However, given the limited sample size, the analysis is probably lacking statistical power. Thus, we evaluated if the direction of effects was consistent. Four of the five identified circRNAs, circ*AFF2*, circ*ETFA*, circ*SPI1* and circ*SUZ12*, were consistent in their effect direction, which suggests that the findings are not ancestry dependent (Supplemental Table 4).

### CircRNA accumulation corelates with symptom severity

We evaluated if any of the identified circRNA were correlated with PD severity measured by UPDRS-III, or neurocognitive decline measured by MoCA (Table 1). We considered correlations significant if nominal p-value was lower than 0.05. Circ*SPI1* showed a significant correlation with UPDRS-III (p=1.45×10^-7^; r^2^=0.015), along with circ*AFF2* following closely (p=1.14×10^-6^; r^2^=0.013). Interestingly, circ*ITGAX* (p=2.81×10^-8^; r^2^=0.017), circ*NCF1* (p=3.53×10^-9^; r^2^=0.019) and circ*PADI4* (p=1.14×10^-8^; r^2^=0.018), which were found to be associated with medication, also showed significant correlation with UPDRS-III. Regarding cognitive measurements, we found that circ*AFF2* was significantly correlated with MoCA (r^2^=0.003; p=0.031), again, with a moderate correlation value (Supplemental Table 6).

### CircRNA accumulation does not reflect disease-associated changes over time in individuals with Parkinson’s Disease

Given the longitudinal design of both the PDBP and PPMI studies, we have also investigated circRNAs using mixed models to identify circular transcripts that change over time. We included 547 PDBP participants (N_casses_=376, N_controls_=171) and 617 PPMI participants (N_casses_=474, N_controls_=143) with at least two visits available for each participant (Table 2) and then performed the meta-analysis. When evaluating the intercepts of the mixed model, they were correlated with what we identified in the cross-sectional analysis (Supplemental Figure 3, Supplemental Table 7), supporting our findings that there are changes in circRNA landscape associated with the presence of the disease. When evaluating significance for the five circRNAs identified in the cross-sectional study, none of them remained significant in their interaction with time. However, all five were consistent in the direction of effect in the interaction term compared to cross-sectional results (Figure 1B, Supplemental Table7). Overall, given the fact that the intercepts were found significant, it suggests that the levels of the five circRNAs do not change over time but due to the presence of the disease. In the meta-analysis, we identified 170 circRNAs with significantly different trajectories between PD and cases. The three most significant findings in the interaction term were circ*GPBP1L1* (beta=0.059, p= 1.351×10^-4^), circ*LAMP1* (beta=0.020, p= 5.467×10^-4^) and circ*C1GALT1* (beta=-0.099, p= 6.859×10^-4^) (Supplemental Table 7).

### Mutations in *LRRK2*, *GBA*, and *SNCA* influence the circRNA landscape

We investigated if there were differences between mutation carriers and sporadic PD cases in the accumulation of the five circRNAs identified in the cross-sectional analysis, in other words, if mutations were contributing to the association. We found that three of the five circRNAs, circ*AFF2*, circ*SUZ12*, and circ*SPI1*, were differentially accumulated in both sporadic PD and mutation carriers (Supplemental Table 4), though their expression was significantly impacted by mutation status (circ*AFF2* log_2_FC=0.286, p=1.752×10^-6^; circ*SUZ12* log_2_FC=-0.258, p= 7.106×10^-3^; circ*SPI1* log_2_FC=0.309, p=6.699×10^-7^). In contrast, circ*FAM13B* was differentially accumulated in mutation carriers (log_2_FC=-0.577, p=6.238×10^-11^) but not in sporadic PD, suggesting that the signal identified in our previous analysis was driven by the familial form of the disease. The opposite was true of circ*ETFA* which was significant in sporadic PD (log_2_FC=-0.190, p=0.002) and not in mutation carriers (Supplemental Table 4). When broken down by mutation, circ*FAM13B* was differentially accumulated in both *LRRK2*^+^ (log_2_FC=0.252, p=0.003) and *GBA*^+^ (log_2_FC=−0.749, p=4.370×10^-8^), whereas circ*AFF2* (log_2_FC=0.776, p=1.467×10^-4^) was differentially accumulated in *SNCA*^+^ carriers only (Supplemental Table 4). Altogether, suggesting the presence of heterogeneity in the circular transcriptome in relation to PD genetic background.

We followed the same approach using the longitudinal data. None of the five circRNAs were significant in the interaction term between disease status and time in sporadic PD participants, but one, cir*AFF2* (beta=0.085, p=6.955×10^-3^), was significant (Supplemental Table 8) in mutation carriers. Direction of effect was largely consistent with previous observations in both sporadic and familial cases, with the exception of circ*ETFA* which had opposite effect in mutation carriers (beta=0.030, p=0.411; Supplemental Table 8). Divided by gene, two out of five circRNAs, circ*AFF2* (beta=0.072, p=0.036) and circ*FAM13B* (beta=−0.106, p=0.028) were associated with the interaction term in *LRRK2*^+^ mutation carriers, and circ*AFF2* (beta=0.341, p=0.015) in *GBA*^+^ carriers (Supplemental Table 8), suggesting that the circ*AFF2* association might be driven by the *GBA*^+^ carriers. Longitudinal analyses were not possible for SNCA^+^ carriers due to very limited sample size (N=8).

### circRNA accumulation starts before symptom onset

We examined whether changes in circRNA accumulation can be observed in participants who are at high risk of developing PD, namely carriers of known PD-related mutations that have not been diagnosed, or those who show known PD associated syndromes like, RBD or hyposmia. These analyses were done in the PPMI dataset exclusively due to prodromal participant availability, and included 150 *LRRK2*^+^, 87 *GBA*^+^, and three *SNCA*^+^ mutation carriers, along with 25 participants exhibiting RBD, and 18 participants with hyposmia. Similar to symptomatic participants, we found that circ*AFF2*, circ*FAM13B*, circ*SUZ12* and circ*SPI1*, were differentially expressed in *LRRK2*^+^ (circ*AFF2*, log_2_FC=0.294, p=1.42×10^-4^; circ*FAM13B*, log_2_FC=-0.389, p=3.80×10^-5^; circ*SUZ12*, log_2_FC=-0.476, p=7.09×10^-5^; circ*SPI1*, log_2_FC=0.348, p=5.01×10^-5^) and *GBA*^+^ (circ*AFF2*, log_2_FC=0.270, p=0.002; circ*FAM13B*, log_2_FC=-0.548, p=4.38×10^-7^; circ*SUZ12*, log_2_FC=-0.714, p=1.73×10^-6^; circ*SPI1*, log_2_FC=0.321, p=0.001) carriers compared to healthy controls (Supplemental Table 9). Additionally, circ*ETFA* (log_2_FC=−0.177, p=0.045) was associated with *GBA*^+^ carriers (Supplemental Table9). Further, we found four circRNAs, circ*AFF2* (log_2_FC=0.507, p=4.030×10^-4^), circ*FAM13B* (log_2_FC=-0.477, p=0.006), circ*SPI1* (log_2_FC=0.562, p=5.260×10^-4^) and circ*SUZ12* (log_2_FC=-0.469, p=0.048), differentially expressed in participants with RBD, and three circRNAs, circ*AFF2* (log_2_FC=0.317, p=0.046), circ*FAM13B* (log_2_FC=-0.406, p=0.026) and circ*SPI1* (log_2_FC=0.428, p=0.023), differentially accumulated in participants with hyposmia.

Next, we compared circRNA expression between at risk and symptomatic participants and found that two circRNAs, circ*AFF2* (log_2_FC=0.178, p=0.022) and circ*SPI1* (log_2_FC=0.168, p=0.038), were associated with disease status in symptomatic compared to at risk *LRRK2*^+^ carriers and one, circ*AFF2* (log_2_FC=0.299, p=0.011), in *GBA*^+^ carriers (Supplemental Table 9), suggesting that the cirRNA changes occur prior to symptom onset.

### Targeted miRNAs analysis suggests involvement of circRNA in known PD related pathways

To better understand the biological implications of the five circRNAs identified in the previous analysis, we listed their predicted miRNA targets using CircInteractome web tool^28^. By doing so, we obtained a list of 282 miRNA targets for circ*AFF2*, 96 for circ*ETFA*, 178 for circ*FAM13B*, 26 for circ*SPI1,* and 313 for circ*SUZ12*. To ensure the biological relevance of the analyses and reduce the number of miRNAs, we leveraged the miRNA counts available in the PPMI dataset. We performed correlation analyses between normalized circRNA and normalized miRNA counts to retain the miRNA binding sites with biological evidence (p<0.05 for the correlation between circRNA and miRNA) for downstream analyses. We reduced the miRNA targets to 27 miRNAs for circ*AFF2*, 7 miRNAs for circ*ETFA*, 20 miRNAs for circ*FAM13B*, two miRNAs for circ*SPI1*, and 30 miRNAs for circ*SUZ12* (Supplemental Table10). We then performed pathway analyses with DIANA mirPath using the 61 miRNAs as input. We observed enrichment in several KEGG terms such as dopaminergic synapse^34–36^ (p=5.411×10^-4^) and long-term depression^37^ (p= 6.20×10^-5^), both of them previously associated with PD, and enriched with miRNAs associated with four of the five circRNAs (Supplemental Table11). Another term that we found enriched in miRNAs correlated with all five circRNAs is the Hippo signaling pathway (p=7.68×10^-8^) (Supplemental Table 11), which has previously been described to play a role in ischemia-associated CNS diseases and PD^38,39^. Ubiquitin mediated proteolysis, suspected to be impaired in PD and contributing to Lewy body formation^40–42^, was also found enriched (p=6.32×10^-3^) for miRNAs targeted by three of the five circRNAs (Supplemental Table 11). Together, these findings add evidence to the potentially role of circRNAs in PD pathogenesis.

### CircRNAs are informative as biomarkers of early stages of Parkinson’s disease to potentially aid clinical diagnosis

To test whether circRNA levels can be used to differentiate between early-stage PD and healthy controls, we developed and evaluated several predictive models. We included first visits from both PDBP (N= 528) and PPMI (N=617) datasets. We evaluated two linear, L1 and L2 regularization, and two non-linear, random forests and MiniPatch, machine learning approaches. MiniPatch model with 149 circRNAs performed best (AUC=0.825; Supplemental Figure 5), followed by L2 regularization with 227 circRNAs (AUC=0.784; Supplemental Figure 5). Interestingly, 51 circRNAs are shared between the two models.

## DISCUSSION

In this study we leveraged publicly available longitudinal blood RNAseq data from two of the largest PD studies to date, PDBP and PPMI, to identify circRNAs that were differentially accumulated in relation to PD. Recent studies have also investigated circRNA differences in brain and blood of PD patients with interesting results, though limited in power.^13,14^ To our knowledge, the present study is the largest to date (N=1,848 subjects, and N=4,833 samples), consisting of two large studies, PDBP, and PPMI, with samples and data collected consistently for each of them. Additionally, the design of the cross-sectional analysis attempted to maximize differences between cases and controls by including the last visit of each individual instead of baseline as previous reports.^14^ By including the last visit, the disease is more advanced, and thus the differences are potentially more pronounced. Finally, and for the first time in PD, we have taken into consideration the longitudinal design of the study and included multiple observations per participant to not only identify circRNA that are potentially differentially accumulated but also find those that change with time and have the potential to be leveraged to follow the progression of the disease.

Overall, we identified five high-confidence circRNA transcripts, circ*AFF2*, circ*ETFA*, circ*SPI1*, circ*SUZ12*, and circ*FAM13B*, that were associated with PD at the time of the last visit whose association was not caused by cellular composition differences or medication use. Given the longitudinal nature of the data, we also evaluated how the five circRNAs related with the advancement of the disease. Despite the lack of statistically significant association between at the interaction term level, all direction of effects were consistent between cross-sectional and longitudinal suggesting that the mixed model might be underpowered, and the circRNAs might potentially change with disease. When contextualizing biologically he findings by predicting miRNA binding sites, we observed an enrichment in dopaminergic synapse term. It is well established that PD is caused by the death of dopaminergic neurons^34–36^, thus, it is plausible to think that circRNAs might be participating as regulatory molecules. Strikingly, the neuronal death happens in brain, but we are capturing the signal in blood, suggesting that those changes are being captured as the result of blood-brain barrier disruption.^16^ Additionally, we found Hippo pathway^38,39^, which has already been linked with PD, to be enriched in conjunction to all our circRNAs. Together with enrichment in other PD-associated pathways such as long-term depression and ubiquitin-mediated proteolysis, these results show that blood circRNAs reflect changes related to PD pathobiology, potentially leaking from the brain. Finally, the circRNA not only capture relevant biological events, but we have leveraging 149 of them to accurately differentiate PD from healthy controls, with an AUC of 0.825.

We further evaluated our findings in diverse genetic backgrounds, finding that both ancestry and mutation carrier status contributed to PD circRNA landscape. Though our results point to shared circRNA expression patterns between participants of African and those of European ancestry, there is an urgent need to increase the diversity of the cohorts to not only validate our findings, and those from others, but also to understand better the pathobiology underlying PD.

*FAM13B* was found to be differentially expressed in PD brains but not in blood.^43^ Interestingly, another study including eight participants did report the dysregulation of *FAM13B* in blood.^13^ In the present study with more than 1,800 subjects, we observed changes in *FAM13B* in blood of PD participants, confirming that *FAM13B* is indeed differentially accumulated in the blood. Additionally, we also report that the circular form of *FAM13B* is differentially accumulated in blood of PD participants compared to controls. Overall, these findings suggest that *FAM13B* is a key player in PD pathogenesis. The presence of circular forms suggests that there are tightly regulated processes associated with this gene. Looking at longitudinal data, circ*FAM13B* was significant in the disease:time interaction term in *LRRK2*^+^ carriers, but not other participants with the familial form of PD. This could indicate that while circ*FAM13B* does correlate with PD diagnosis in all mutation carriers, it does so via a different process in *LRRK2*^+^ which more closely reflects the advancement of the disease.

*SPI1*, a transcription factor involved in myeloid cell development and function, is known for its role in Alzheimer’s disease (AD).^44^ Given its association with AD, and our findings, *circSPI1* was found associated with PD independently of medication and cell counts, we investigated if the association with PD status might be driven by cognitive function measured by MoCA. Unfortunately, the correlation was not significant. Despite the negative results and given the close to normal values of MoCA in the population included in this study, it is plausible to think that this correlation might become significant once the disease advances. Alternatively, circ*SPI1* might influence PD differently than AD, or even have a different function than the linear form, thus explaining the lack of association.

Among the novel circRNA associations described in this paper, several of the host genes have not been previously linked to PD. For example, *AFF2* hasn’t been previously reported in PD, but it has been reported to contribute to axonal degeneration and TDP-43 pathology in frontotemporal dementia (FTD) and amyotrophic lateral sclerosis (ALS).^45^ Given the involvement of *AFF2* in neurodevelopment^46,47^ and neurodegeneration^45^, it would be reasonable to postulate that circ*AFF2* is integral to proper functioning of the nervous system as well. This is further supported by our finding of circ*AFF2* being significantly differentially expressed, including across different genetic backgrounds, with consistent direction of effect.

Regarding PD-related mutation, this study included participants with mutations in *LRRK2*, *GBA* and *SNCA* genes. The most common *LRRK2* mutation, Gly2019Ser, leads to constitutive activation of *LRRK2*, which leads to activation of neuronal death pathway, and possibly upregulation of *SNCA*.^48,49^ On the other hand, the most common *SNCA* mutations are copy-number mutation, more specifically duplications.^50^ Altogether, it is not surprising to observe such heterogeneity in circRNA landscape, as regulatory pathways that are triggered by *LRRK2* mutation to bring about PD, might not be affected in *SNCA* mutation carriers, given that *SNCA* is downstream from *LRRK2*. Our findings regarding circ*FAM13B*, circ*SPI1*, and circ*SUZ12* that are significantly associated with *LRRK2* mutation carriers, but not *SNCA* seem to support this hypothesis. Differences in pathway regulation between *LRRK2* and *SNCA* mutation carriers are further emphasized by opposing directions of effect of circ*ETFA* in the two. Mutations in *GBA* gene lead to decrease in GCase enzyme activity, resulting in lysosome malfunctioning. GCAse impairment has been suggested to promote alpha-synuclein accumulation in PD. This points to possible overlap between regulatory pathways, that involve circ*FAM13B*, circ*SPI1*, and circ*SUZ12*, that are differentially accumulated in *GBA* and *LRRK2* mutation carriers, potentially leading to increased alpha-synuclein pathology in both. Further analyses in a larger sample would be required to verify these findings.

This study had several limitations. While we had access to the two largest longitudinal datasets (4,833 samples, 1,789 circRNAs), longitudinal analyses are underpowered, and larger sample sizes are needed to appropriately power these analyses. Likewise, the number of African American participants, at-risk individuals, or mutation carriers was very limited, thus we did not perform *de novo* discovery in these groups, but rather leveraged them to validate the main findings. Further efforts are needed to actively recruit participants from diverse backgrounds. Furthermore, mutation and medication data were absent from the PDBP dataset, along with the recruitment of “at risk” individuals, which did not allow for a straight comparison of the two populations. Finally, we are repurposing traditional RNAseq data to identify and quantify circRNAs, rather than purifying and subsequently sequencing circRNAs. Despite the need for some additional analysis and validation in the future, we have successfully replicated findings from other groups, supporting the validity of this approach.

In conclusion, this is the largest study to date describing and biologically contextualizing the circRNA landscape in blood in relation to PD. We identified and replicated five circRNAs differentially accumulated in PD compared to healthy controls and linked to biologically pathways relevant for the disease. Despite the limitations, we have performed several sensitivity analyses to account for ethnic and disease diversity demonstrating that circRNA not only have a biological role in PD but can also be leveraged as biomarkers to potentially aid in the clinical diagnosis.

## Data Availability

All data are openly available from the PDBP and PPMI studies

https://www.ppmi-info.org/access-data-specimens/download-data

https://www.amp-pd.org/register-for-amp-pd

## Acknowledgments

We thank all the participants and their families along with the institutions and all the staff who provided plasma tissue, without whom this study would not have been possible. We thank Dr. Abdallah Eteleeb for providing an outline upon which we built our data processing pipeline and for his hands-on help processing part of the data. Next, we would like to thank Drs. Joe Boktor and Sarkis Mazmanian, for kindly providing the deconvoluted blood cell counts using Immunedeconv tool that was leveraged to add cellular composition in the present manuscript. This work was supported by access to equipment made possible by the Hope Center for Neurological Disorders, the Neurogenomics and Informatics Center (NGI: https://neurogenomics.wustl.edu/) and the Departments of Neurology and Psychiatry at Washington University School of Medicine.

## Author Contributions

LI, AB and YS conceived and wrote this article. LI and CC conceptualized the research plan. LI, AB, and YS designed the analysis plan. YS, AB, SS, TP, JS, and CM performed the analyses. JAP and JB performed predictive modeling. AB, YS, SS, CM, JAP, JB, CC, and LI discussed the project, revised the manuscript, and provided critical feedback.

