## SupplementalFigures for "Circulating blood circular RNA in Parkinson’s Disease; a systematic study"

**Correspondence:**

Laura Ibanez, PhD

**Supplemental Figure 1:** Quality control summary for circular RNA count processing. Upset plots indicate number of circRNAs common across visits in **A.** PDBP, and **B.** PPMI. Only those common across all visits and present in both studies were kept for analyses. Scatter plots show the principal components for **C.** PDBP and **D.** PPMI calculated using the 500 most variable circRNA transcripts prior to quality control. Individuals outside three-standard deviation (marked with the dashed square) were removed from further analyses.

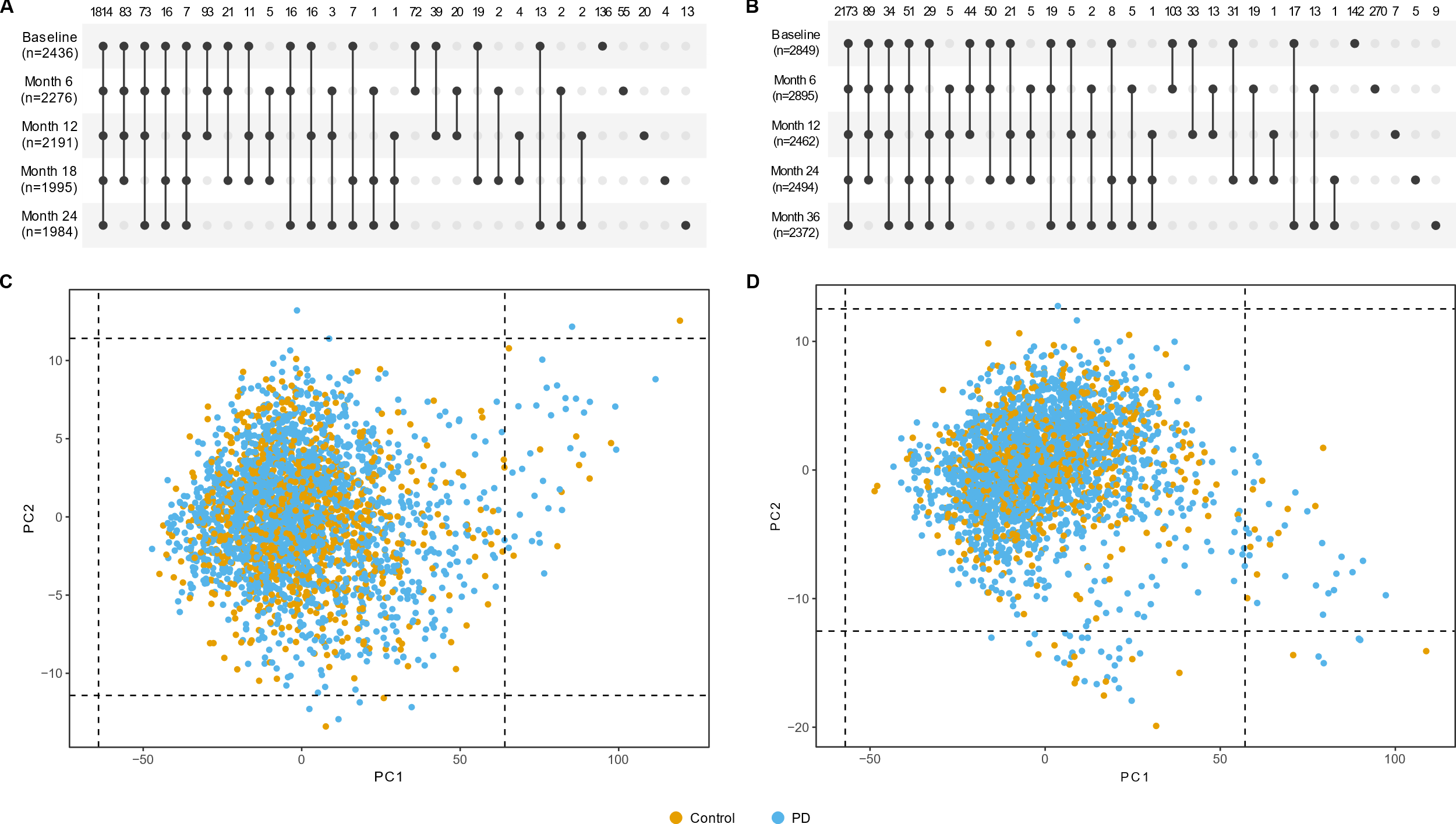

**Supplemental Figure 2.** Violin plots showing levels of the three circRNAs that associated with medication; **A.** *circTGAX;* **B.** *circNCF1;* and **C.** *circPADI4.* Color coded is the normalized count distribution for non-medicated PD cases, yellow, medicated PD cases, brown, and controls, light brown. Asterisks denote p-value: **=0.01, ***=0.001

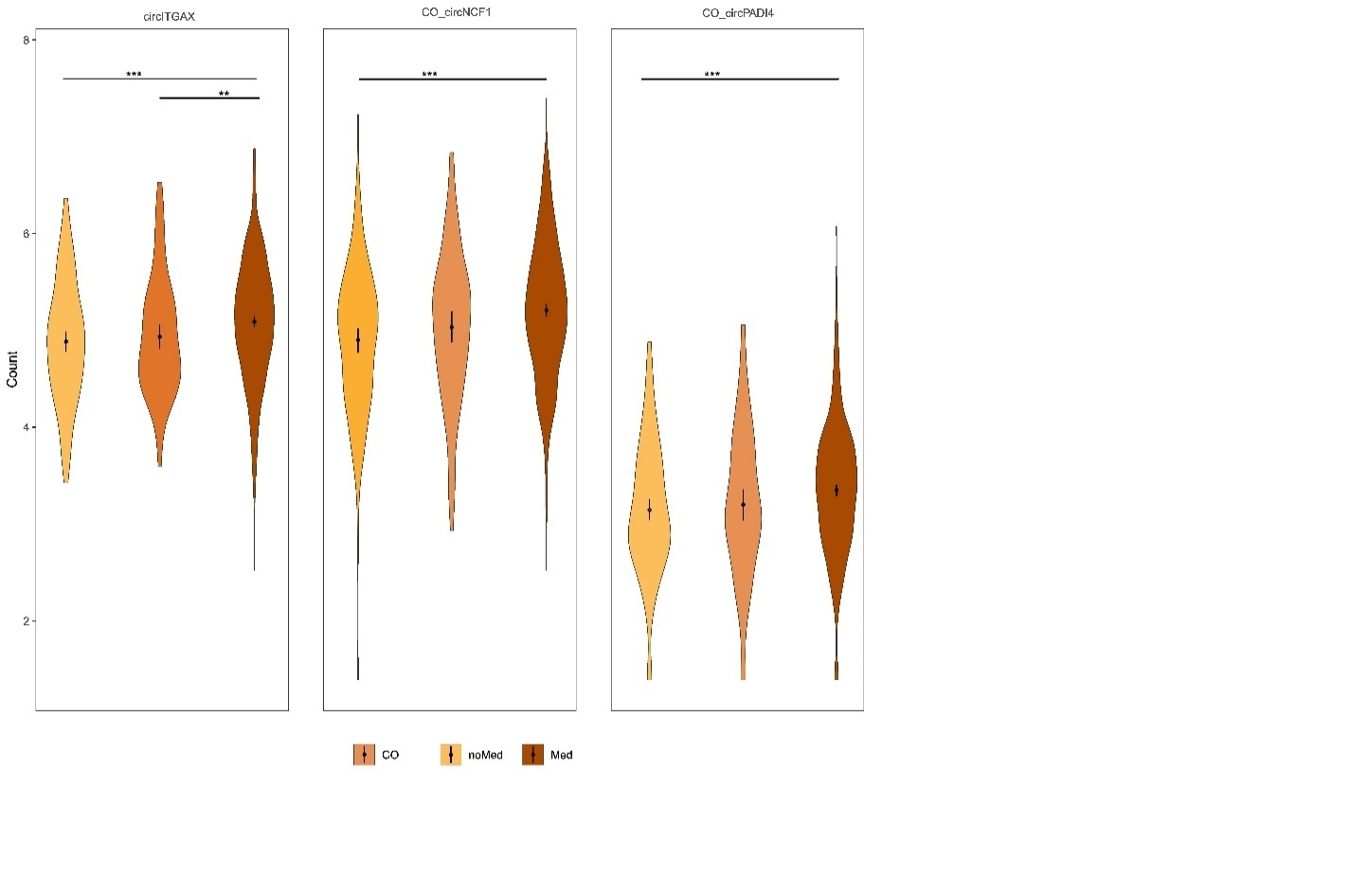

**Supplemental Figure 3.** Scatter plot of significant (p-value<10^-3^) cross-sectional and longitudinal analyses effect sizes in PDBP dataset; due to the nature of the data (normalized transcript counts), all intercepts from the mixed model were positive, so absolute values were used for cross-sectional effect sizes

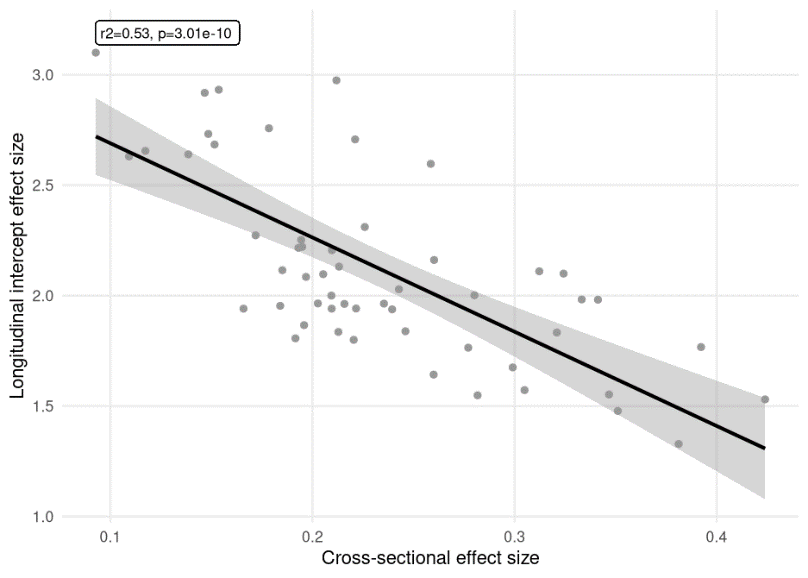

**Supplemental Figure 4.** Violin plots representing the levels of the five differentially expressed circRNAs (circ*AFF2*, circ*ETFA*, circ*FAM13B*, circ*SPI1*, and circ*SUZ12*) in different subgroups of the PPMI population. **A.** Heathy controls (brown), at risk (yellow), and symptomatic (grey) participants, **B.** at risk participants broken down by mutation carrier status, and **C.** symptomatic participants broken down by mutation carrier status; asterisks denote p-value: *=0.05, **=0.01, ***=0.001

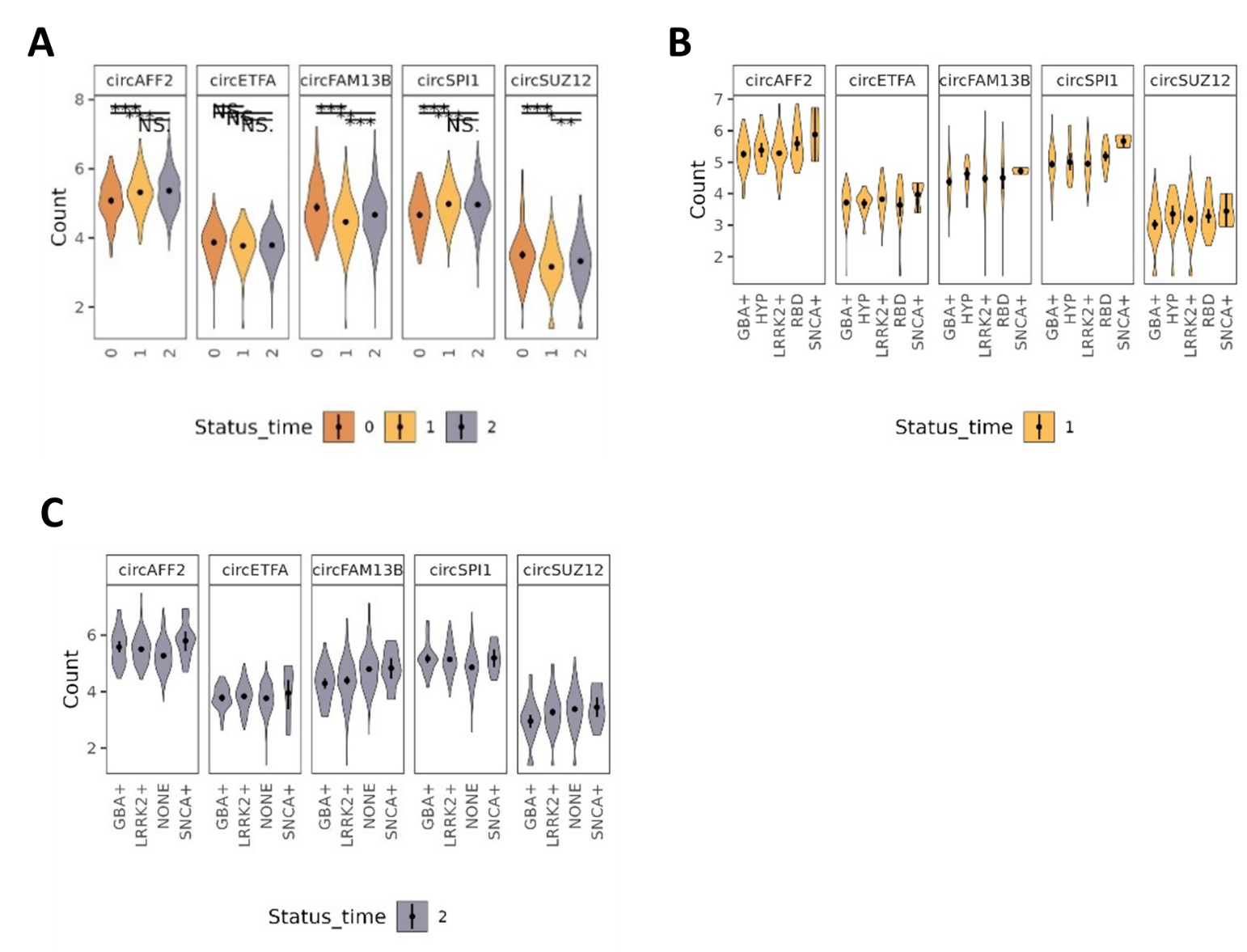

**Supplemental Figure 5.** ROC curves for the best performing Ridge (purple), LASSO (blue), Ranger (yellow) and MiniPatch (pink) predictive models in the testing (PPMI) population
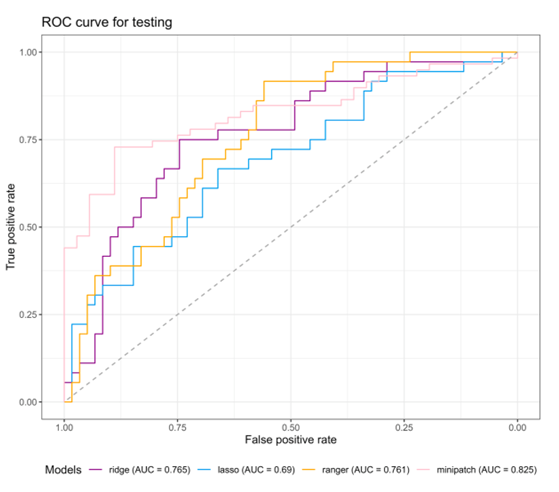
